# Rare *CRHR2* and *GRM8* variants identified as candidate factors associated with eating disorders in Japanese patients

**DOI:** 10.1101/2023.08.24.23294455

**Authors:** Akira Oka, Shinji Hadano, Mahoko Takahashi Ueda, So Nakagawa, Gen Komaki, Tetsuya Ando

## Abstract

Eating disorders (EDs) are a type of psychiatric disorder characterized by pathological eating and related behavior and considered to be highly heritable. The purpose of this study was to explore rare variants expected to display biological functions associated with the etiology of EDs. We performed whole exome sequencing (WES) of affected sib-pairs corresponding to disease subtype through their lifetime and their parents. From those results, rare single nucleotide variants (SNVs) concordant with sib-pairs were extracted and estimated to be most deleterious in the examined families. Two non-synonymous SNVs located on corticotropin releasing hormone receptor 2 (*CRHR2*) and glutamate metabotropic receptor 8 (*GRM8*) were identified as candidate disease susceptibility factors. The SNV of *CRHR2* was included within the cholesterol binding motif of the transmembrane helices region, while the SNV of *GRM8* was found to contribute to hydrogen bonds for an α-helix structure. CRHR2 plays important roles in the serotoninergic system of dorsal raphe nuclei, which is involved with feeding and stress-coping behavior. Moreover, GRM8 modulates glutamatergic neurotransmission, and is also considered to have effects on dopaminergic and adrenergic neurotransmission. Further investigation regarding the biological function of these variants may provide an opportunity for elucidate the pathogenesis of EDs.

## 1. Introduction

Eating behavior can be severely affected by eating disorders (EDs), including anorexia nervosa (AN) and bulimia nervosa (BN). The main manifestations of AN are dietary restriction, fasting, excessive exercise, or other weight control or loss behaviors as resistance to maintenance of normal body weight. On the other hand, BN is characterized by repeated binge eating and inadequate compensatory behaviors to avoid weight gain. Manifestations shared by these disorders include aspiration for weight loss, fear of weight gain, and self-evaluation unduly influenced by body shape and weight. Moreover, twin and familial studies demonstrated that genetic predisposition to AN and BN is shared, at least in part [1–3]. The Diagnostic and Statistical Manual of Mental Disorders (DSM-5) provides diagnostic criteria for a subgroup of AN defined as restricting type (AN-R), which include severe dietary restriction without regular binge eating and/or purging behaviors, as well as binge-eating/purging type (AN-BP) with regular binge eating and/or purging behaviors.

A recent GWAS for AN in European identified eight significant common variants [4]. However, that study captured only 1.7% of the phenotypic variance, thus additional subjects would be required for future replication studies [4]. On the other hand, a sequencing study with linkage analysis was performed using two large families with AN and BN, and those findings suggested that each rare missense variant in estrogen-related receptor α (*ESRRA*) and histone deacetylase 4 (*HDAC4*) is associated with EDs [3]. *ESRRA* null mice also display behavioral deficits relevant to EDs such as AN [5]. Moreover, *HDAC4*^A778T^ mice carrying the missense variant demonstrate several ED-related feeding and behavioral deficits in a gender-dependent manner [6, 7].

Swedish twin study for EDs demonstrated that concordance rates in monozygotic twins were higher than dizygotic twins within the same disease subtype, on the other hand, the rates were similar in the different subtype [1]. Therefore, in the current study, to use subjects expected to be higher heritability we selected seven affected sib-pairs corresponding to disease subtype through their lifetime for whole exome sequencing (WES) for exploring rare variants functionally related to the pathogenesis of EDs. However, the statistical power was quite insufficient to perform genetic association and linkage analysis. Therefore, we explored the most deleterious coding variants shared with sib-pairs in each family.

## 2. Materials and methods

### 2.1. Patients and families

Upon approval of the experimental procedures from the relevant ethical committees at the National Institute of Mental Health, National Center of Neurology and Psychiatry (number: A2013-054) and Tokai University (number: 14I-60), we obtained written informed consent in accordance with the Declaration of Helsinki from 30 sib pairs and their family members prior to collection of DNA samples. Among all subjects we selected 7 sib pairs corresponding to disease subtype through their lifetime for reduction of sample heterogeneity and elevation of likelihood of identification of susceptibility variants [8], though family P8 included a brother who displayed a different subtype (**Fig. 1 and Supplementary Table 1**). The IDs of the samples in this paper were reassigned for the experiment, therefore these IDs could not identify individuals. They are 15 individuals affected with EDs and 11 unaffected individuals, all of Japanese origin. They are also three ANR and three BN families, and one family displayed a diagnostic crossover from ANR to BN. Of the cases, each was diagnosed according to the DSM-IV criteria in Japan. DSV-IV eating disorder diagnosis was made by expert of psychosomatic doctors or psychiatrists expertized on eating disorders. DNA was extracted using a QIAamp DNA blood kit (QIAGEN, Hilden, Germany).

**FIGURE 1.**
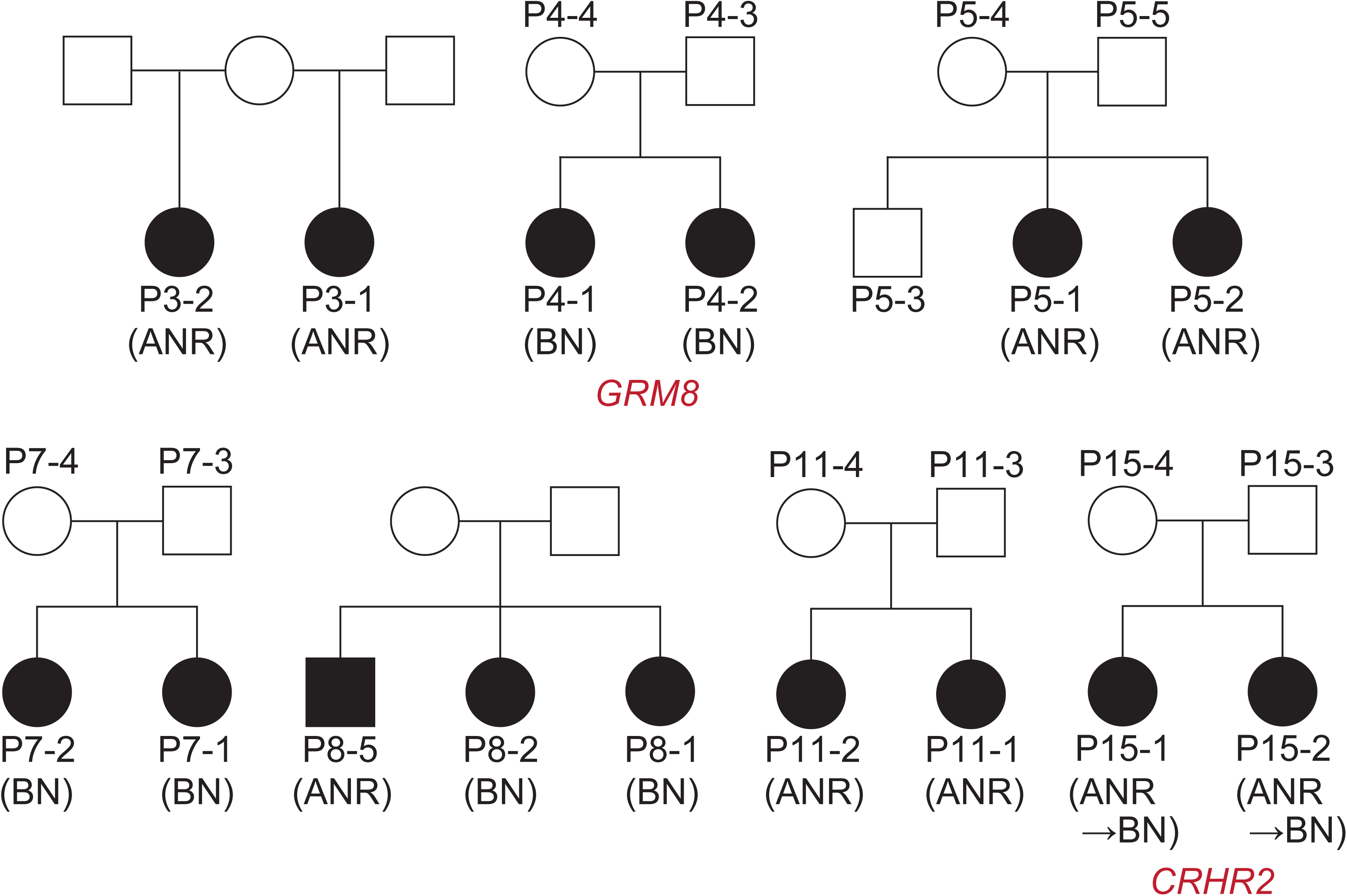
Sib-pair families affected by EDs were examined using whole exome sequencing. Individuals labeled with an ID number were sequenced. The disease phenotype of affected sib-pairs in the P15 family was transferred from ANR to BN. The rare variants *GRM8* and *CRHR2* were observed in the P4 and P15 families, respectively. Abbreviations: ANR, anorexia nervosa restricting type BN, bulimia nervosa

### 2.2. Genomic library construction and sequencing

For exon fragment capture and sequencing, an Agilent SureSelect Target Enrichment, v. 5 (50 Mbp), was used. Sequence analysis was performed using an Illumina Genome Analyzer IIx, HiSeq2000, or HiSeq2500 platform.

### 2.3. Sequencing data analysis

Reads that passed quality control were mapped to the reference genome (UCSC Genome Browser assembly GRCh37/hg19, http://genome.ucsc.edu/) with a Burrows-Wheeler Aligner, v. 0.5.9.

Potential PCR duplicates were flagged with Picard MarkDuplicates, v. 1.88 (http://picard.sourceforge.net/). Genome Analysis Toolkit v. 2.2-8, was used to perform map quality score recalibration and variant detection. Single nucleotide variants (SNVs) and indels were then annotated for functional consequences at the gene using the ANNOVAR. Predictions of variants with risk were also performed using ANNOVAR, with non-synonymous variants (LJB, v. 3.0) employed for the effects on protein function (SIFT, PolyPhen, MutationTaster, Muration Assesor, LRT, FATHMM) and evolutional conservation (GERP++, phyloP, Siphy). Criteria used for deleterious variants in each prediction are shown following:

SIFT (sift): D: deleterious, PolyPhen 2 HDIV: D: Probably damaging, LRT: D: Deleterious, MutationTaster: A (disease_causing_automatic) or D (disease_causing), MutationAssessor: H: high FATHMM: D: deleterious, GERP++ >5, phyloP >2.0, SiPhy >15

### 2.4. Confirmation of SNVs

For PCR and sequencing (Greiner Bio-one) of the SNV of *CRHR2*, forward (CCTGGCAGGGGGAGAAGAGC) and reverse (CCCCAAGCTGCCTCCTGACA) primers, and for the SNV of *GRM8*, forward (GCAACTCCAAGTCATCCATTTTCTTCA) and reverse (TGGAGCGAATTGCTCGGGATT) primers, were used. Sequencing reactions were carried out using a BigDye Terminator Cycle Sequencing kit, v. 3.1 (Thermo Fisher Scientific). Automated electrophoresis was performed with an ABI PRISM 3730 Genetic Analyzer (Thermo Fisher Scientific).

## 3. Results

### 3.1. WES

We obtained rare 4,032 SNVs, 113 splice site variants and 76 insertion/deletion variants (allele frequency <0.001) from a total of 633,030 variants (**Fig. 2**). Initially, we attempted to identify recessive and compound variants concordant with the affected sib-pairs, however none were found among these rare variants. Next, heterozygote variants concordant with affected sib-pairs were extracted as dominant candidates. Finally, we estimated the most deleterious SNV in each family by use of six functional prediction (SIFT, PolyPhen, MutationTaster, Muration Assesor, LRT, FATHMM) and three conservation-based (GERP++, phyloP, Siphy) methods [9].

**FIGURE 2.**
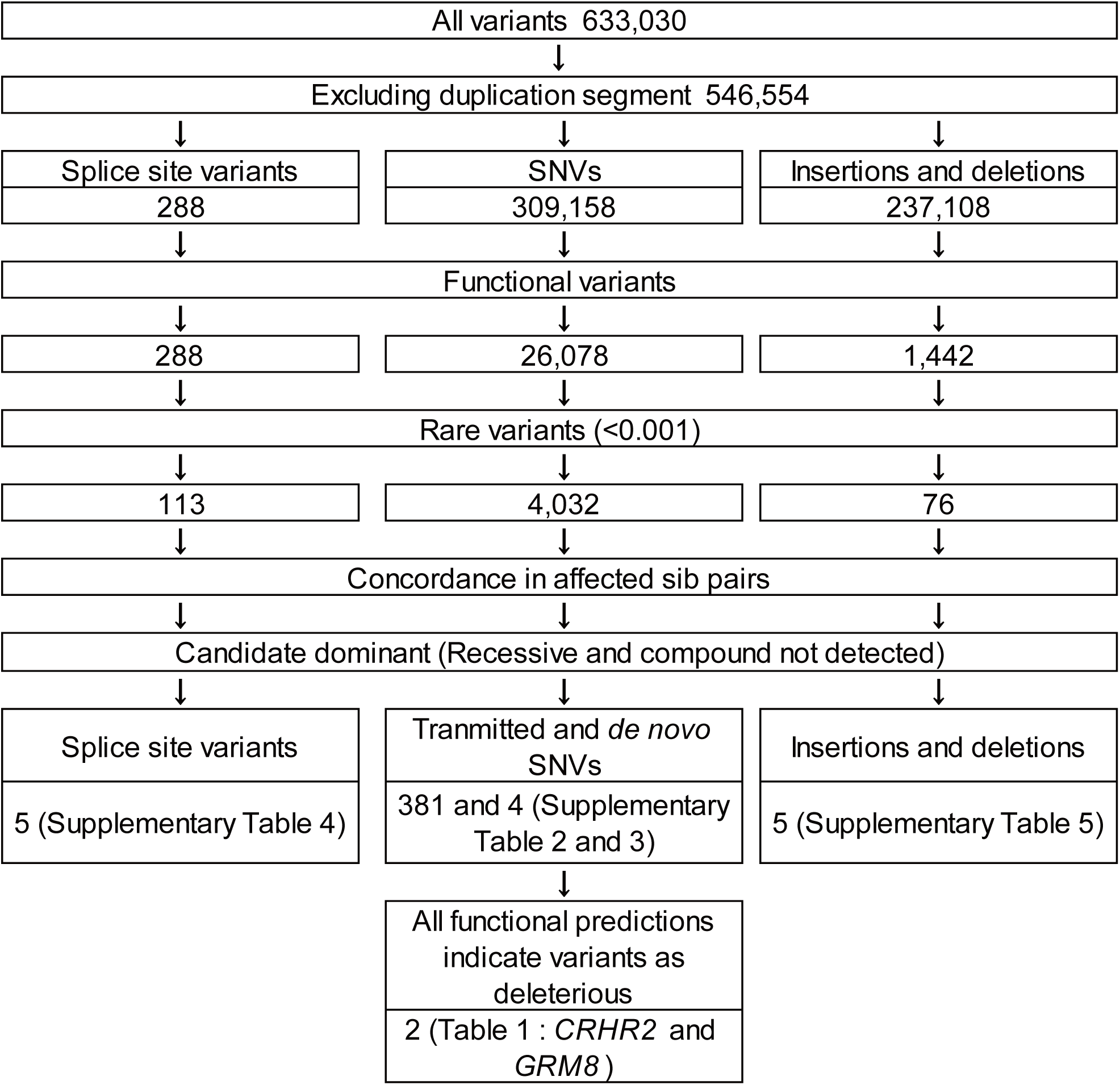
Scheme for screening variants with whole exome sequencing.

A previous large exome sequencing study indicated a strong inverse relationship between average SNV age and number of methods needed to predict a variant as deleterious, thus SNVs predicted to be deleterious by multiple methods probably undergo a more intense purifying selection and are very rare in populations [10], suggesting them as candidates for disease pathogenesis. Moreover, most very-rare variants arose within the past 5–10,000 years and are population-specific [11]. Therefore, 381 SNVs and 4 *de novo* SNVs were evaluated (**Fig. 2, Supplementary Table 2**) based on the number of methods that predicted a variant to be deleterious and a feature of the gene. Among those, only two were predicted to be deleterious by all nine methods, an SNV in corticotropin releasing hormone receptor 2 (*CRHR2*) noted in family P15 and an SNV in glutamate metabotropic receptor 8 (*GRM8*) noted in family P4 (**Table 1 and Fig.1-2**). These were considered likely to be specific for a Japanese population, though a single allele for *CRHR2* was previously observed in Europeans (**Table 1**). In addition, SNVs supported by eight of the methods were observed in the P7, P8, and P11 families, while those supported by seven of the methods were observed in the P3 and P5 families. Among the 10 genes, *CRHR2, GRM8*, and glutamate ionotropic receptor α-amino-3-hydroxy-5-methyl-4-isoxazole propionic acid (AMPA) type subunit 2 (*GRIA2*) were indicated to have higher levels of expression in the brain and related tissues (**Table. 1**).

**Table 1.**
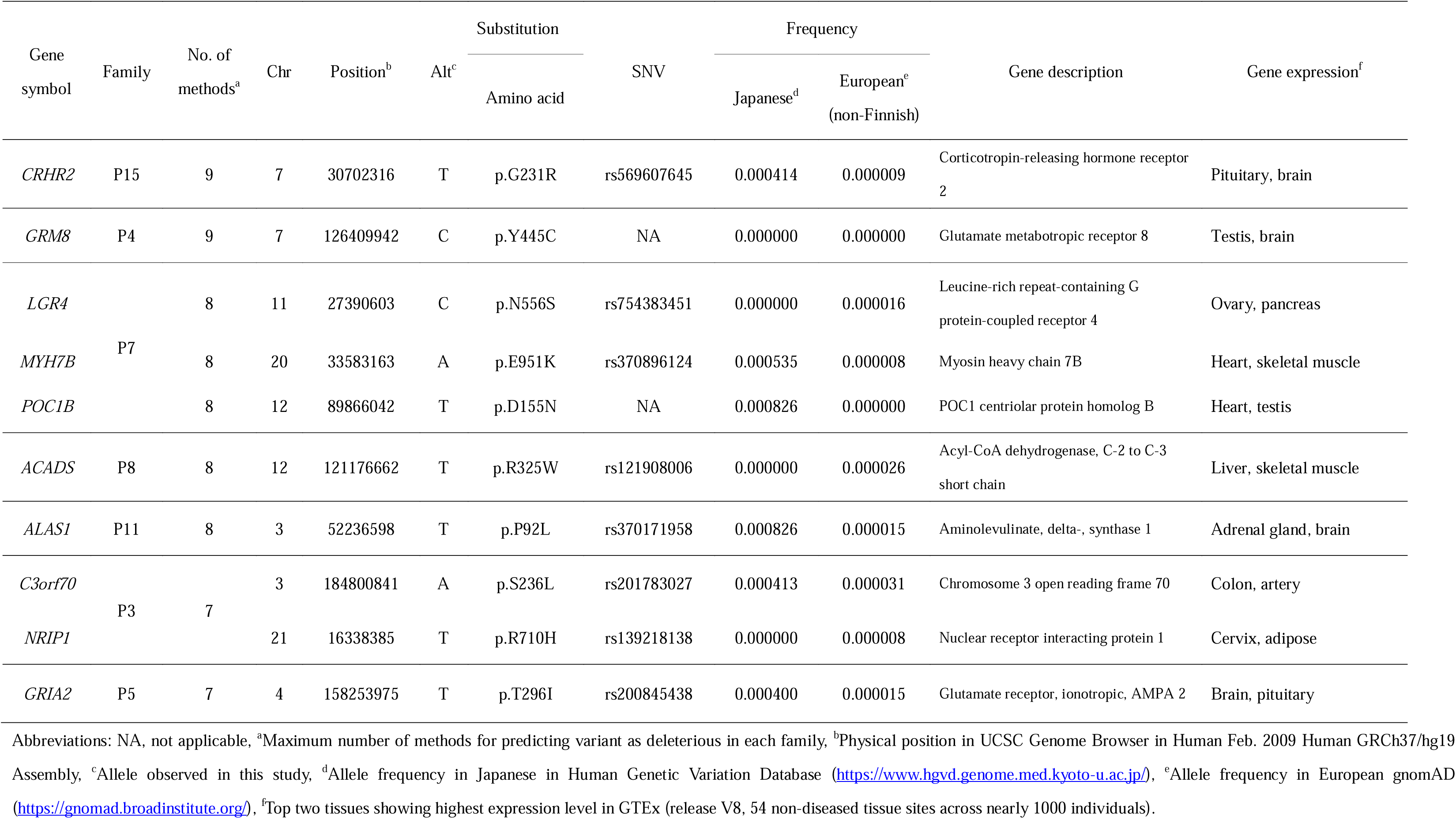
Most deleterious variants in each family estimated by functional prediction.

*CRHR2* encodes a neuropeptide receptor for corticotropin-releasing factor (CRF), a critical coordinator of the hypothalamic-pituitary-adrenal axis [12] and for three urocortins (UCN1–UCN3) [13]. CRHR2 is also a specific receptor for UCN2 and 3, which are associated with regulation of stress, anxiety, and food intake [14–17]. *GRM8* encodes a G-protein coupled metabotropic glutamate receptor that influences inhibition of the cyclic AMP cascade and regulates presynaptic glutamate release [18], and also has been found to have genetic associations with psychiatric phenotypes, including depression [19, 20] and eating behavior [18]. The two variants were also confirmed by Sanger sequencing (**Supplementary Figure 1**). Thus, it is considered that *CRHR2* and *GRM8* are potent candidate genes due to the pathogenesis of EDs in individuals included in this study.

*GRIA2* has a significant role in excitatory neurotransmission [21], and has been identified as a causative gene of intellectual disability and developmental epileptic encephalopathy. Therefore, we considered that *GRIA2* was a weaker candidate gene for EDs in this study, as the phenotypes generated by mutations may not be concordant with EDs. Furthermore, among the 381 SNVs considered to be candidates for dominant, all other SNVs were thought to be unlikely as causative factors of EDs after considering the descriptions and locations of expression **(Table 1)**.

Four *de novo*, five splice site SNVs, and five insertion and deletion variants concordant with affected sib-pairs were identified as heterogeneous (**Supplementary Table 3 - 5**). However, in consideration of their features, these fourteen genes are unlikely to be causative factors of EDs.

### 3.2. Evaluation of impact of SNVs on CRHR2 and GRM8

CRHR2 is classified as a secretin subfamily among class B G protein-coupled receptors (GPCRs). A GPCR features seven transmembrane helices (TM), three extracellular loops, and three intracellular loops. The rare SNV in family P15 (rs569607645) was found located within the TM4 region. Gly 231, with a codon corresponding with the SNV conserved across species and among the class B GPCRs (**Fig. 3**), and located within a highly conserved sequence motif, GWGxP, in class B GPCRs [22]. Moreover, the substitution converts hydrophobic (glycine) into hydrophilic (arginine) amino acid, implying that this SNV influences conformation and function of CRHR2.

**FIGURE 3.**
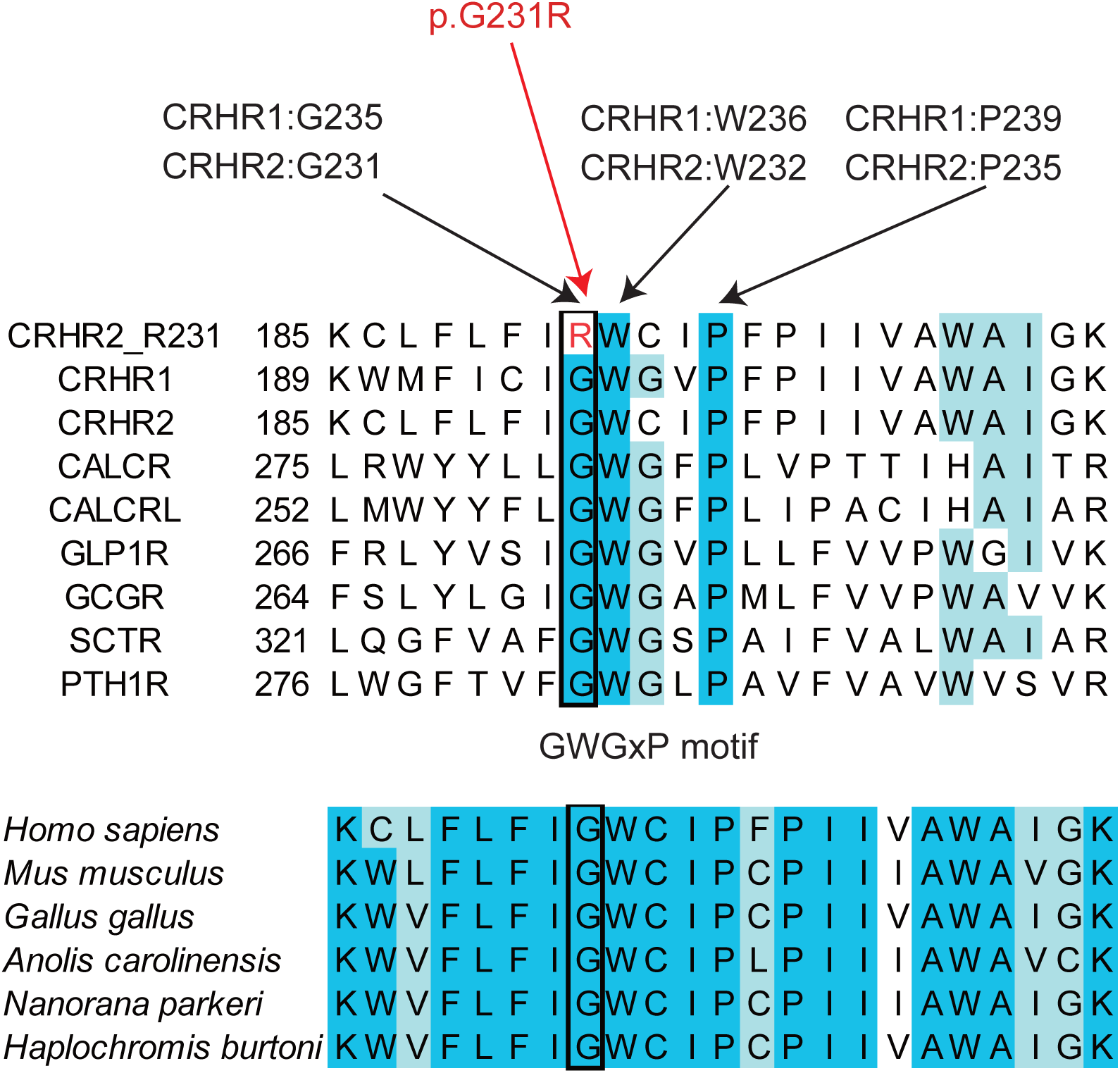
Multiple amino acid sequence alignments of TM4 helix of human class B GPCRs and CRHR2 in various species. Upper alignment indicates TM4 of a selected set of human class B GPCRs included with the GWG x P motif involved in cholesterol binding. Lower alignment indicates evolutionarily conserved amino acids. The level of blue shading is shown according to sequence conservation. Black square indicates amino acid position corresponding to human G231.

Metabotropic glutamate (mGlu) receptors are members of the class C family of GPCRs that modulate cellular responses to the excitatory neurotransmitter L-glutamic acid (L-Glu) in many synapses in the CNS [23]. The amino terminal domain (ATD) of mGlu receptors contains an orthosteric glutamate recognition site, which is highly conserved across the eight mGlu receptor subtypes [24]. The rare SNV in family P4 was located within an α-helix structure of the ATD region. Tyrosine 441, with a codon corresponding with the SNV, was found to be conserved across species and hydrogen bonds to Asp 422 in an adjacent α-helix structure (**Supplementary Figure 2**) [23]. Therefore, the substitution of tryptophan to cysteine is likely to cancel the hydrogen bond. We also evaluated the effect of the SNV on protein stability using X-ray crystal structure data (PDB ID 6BSZ) and ENCoM, which are used to predict the effect of mutations on thermostability and dynamics, as well as to generate geometrically realistic conformational ensembles [25]. Those results indicated that the conformation of GRM8 was expected to be destabilized (-0.822 kcal/mol). Thus, the substitution may have an impact on the structure and functions of GRM8.

## 4. Discussion

### 4.1. CRHR2

Common SNVs in *CRHR2* were not detected in a large genome-wide association study for anorexia nervosa and other psychiatric disorders [4]. On the other hand, WES with bipolar disorder families identified a rare and functionally relevant nonsense variant within the intracellular C-terminal region of *CRHR2* [26]. Thus, it is suggested that a rare SNV in *CRHR2* contributes to the pathogenesis of a psychiatric disorder.

A study that used *Crhr2* knockout mice demonstrated that CRHR2 is essential for sustained feeding suppression induced by UCNs, members of the CRF family of peptides [27]. In another investigation, *Crhr2*-mutant mice showed decreased food intake following food deprivation [12]. Therefore, UCNs are potent for suppression of food intake by CRHR2-specific mediation [28]. Furthermore, *Crhr2*-mutant mice were also reported to have an anxiogenic phenotype and impaired stress recovery [13]. These observations are related to serotoninergic neurons. Serotonin (5-HT) is produced by serotoninergic neurons of the dorsal raphe nucleus (DRN) located within the brain stem and regulates feeding behavior [29], while CRHR2 is specifically expressed in the DRN serotoninergic neuron of [30] and plays important roles in controlling serotonergic neuronal activity [31]. Hammack et al. performed experiments within the rat DRN using UCN2, an agonist for the highly selective CRHR2, as well as an antagonist for that and demonstrated that CRHR2 in the DRN mediates behavioral consequences of uncontrollable stress [32]. Additionally, anxiety-related stimuli by UCN2 were shown to lead to increased activation of serotonergic neurons within the DRN [33]. The DRN projects to the bed nucleus of the stria terminalis, which plays essential roles in threat processing, and is responsible for such emotional states as fear and anxiety [34, 35]. The tone of the DRN–5-HT system is regulated in a dynamic manner through CRHR2 activation, and either decreased or increased depending on the level of endogenous or exogenous ligands [36]. Indeed, in malnourished individuals suffering from AN the cerebrospinal fluid has reduced amounts of 5-hydroxyindoleacetic acid which is the major brain metabolite of 5-HT and is thought to reflect extracellular 5-HT concentrations, indicating that abnormal activity of 5-HT system is related to EDs pathogenesis. [37]. Thus, CRHR2 mediates not only food intake, but also passive coping behavior and depression-like responses triggered by uncontrollable stress [13] via the serotoninergic neuron in the DRN.

Structural analysis demonstrated that the motif included with rs569607645 is a cholesterol-binding site, and that artificial mutations in that site reduce the activation potency of UCN1 and CRF to CRHR2, suggesting that the site plays a role in stabilizing peptide binding [38]. Moreover, bound cholesterols in the structure of CRHR2 contribute to GPCR signaling, indicating that the GPCR signalosome carries out its function in the cholesterol-rich lipid raft [38]. Thus, the substitution of G231R provided by the rare SNV rs569607645 impairs the functions of CRHR2, which may decrease the coping mechanism in response to stress [13] and/or feed suppression [28] in patients with EDs.

ED patients have higher rates of anxiety disorders as compared with normal individuals, suggesting that both disorders might share common etiological factors, which can increase the susceptibility of an individual to either [39]. Stress is prominently involved in the pathogenesis and development of EDs and anxiety disorders. Moreover, coping mechanisms such as a positive attitude, planning, and social support seem to be impaired in ED patients who often do not cope well with emotional distress, implicating that such mechanisms may be disrupted [40]. Therefore, a decay of CRHR2 functions may play a part in the pathogenesis of EDs.

### 4.2. GRM8

As mentioned above, associations between psychiatric disorders and *GRM8* has been indicated with strong evidence in previous reports. On the other hand, a sequencing study for bipolar disorder identified enrichment of damaging mutations in GPCRs including *CRHR2* and the *GRM* gene family, with increased numbers of deleterious variants [26].

The mGluRs modulate glutamatergic neurotransmission, and are considered to also have effects on dopaminergic and adrenergic neurotransmission, implicating their involvement in a number of neurological disorders [41]. An expression study using animals indicated that GRM8 may play a role in feeding behavior and metabolism via the hypothalamic pathway [18, 42].

## 5 Conclusions

We found two rare SNVs as candidates for predisposition to EDs in affected sib-pair families. One found located on *CRHR2* plays important roles in the serotoninergic system in the DRN, and is involved with feeding and stress-coping behavior, while the other SNV is located on *GRM8*, which modulates cellular responses to the excitatory neurotransmitter in the CNS, and may play a role in feeding behavior and metabolism via the hypothalamic pathway. Moreover, the present findings indicated potential for both non-synonymous SNVs to have impact on conformation and functions of molecules. However, this study only observed the deleterious SNVs in one family for each gene and was not able to indicate that the SNVs explained how much of the genetic variance of EDs. Furthermore, it is impossible to rule out all candidate variants. Therefore, these genes must be confirmed by genetic analysis for additional families and sporadic cases and by functional analysis in the future investigations.

## Data accessibility

Data obtained in this study are available from the authors upon reasonable request.

## Author contributions

AO conceived the studies, performed experiments, contributed to the data analysis, and drafted the manuscript. SH, MU and SN were contributed to the data analysis. GK and TA were involved in sample collection, editing the manuscript.

## Data Availability

All data produced in the present study are available upon reasonable request to the authors.

---

**Supplementary Figure 1.**
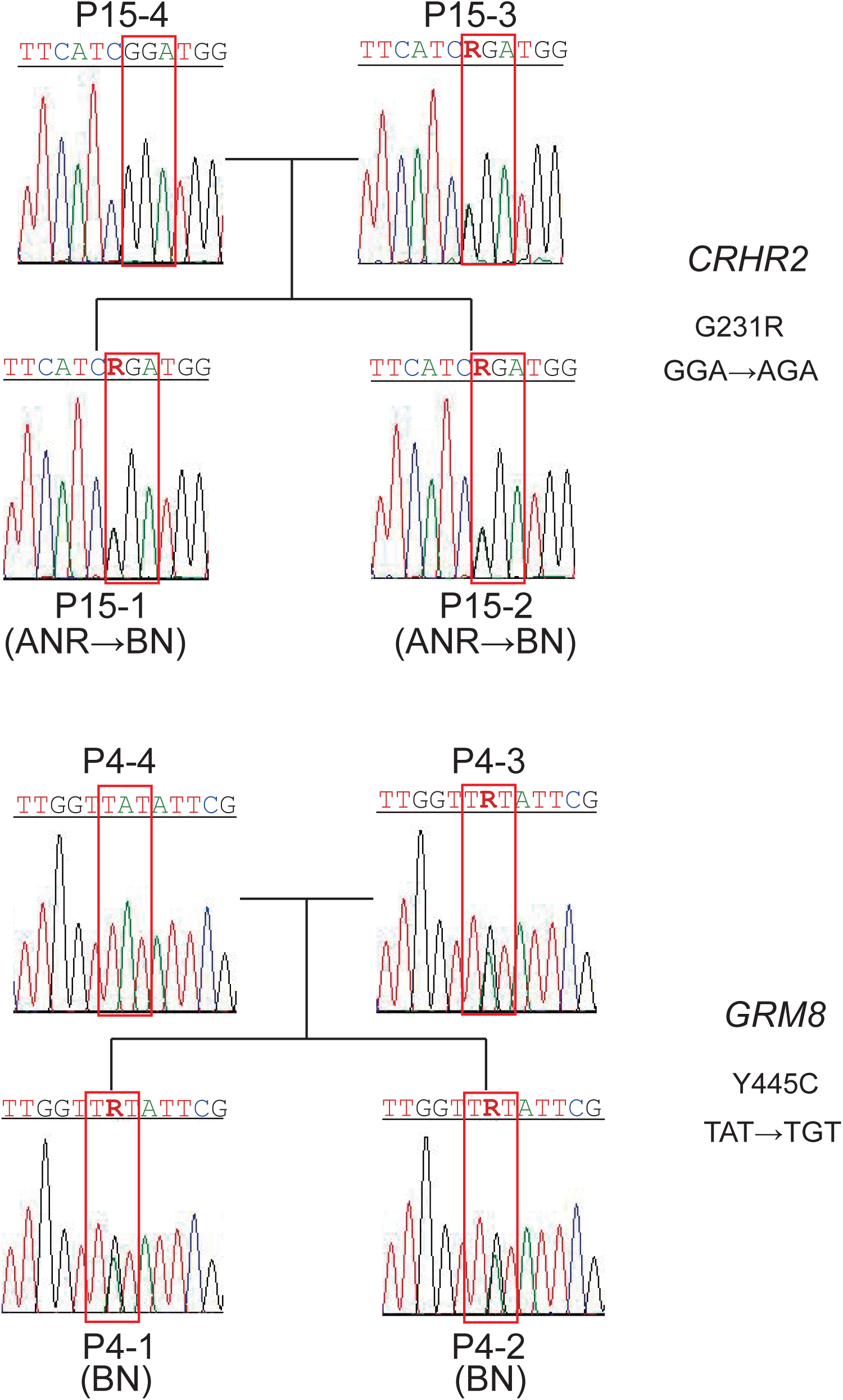
Confirmation by Sanger sequencing for SNVs. Red squares indicate codons providing amino acid substitutions. Upper panel shows SNV of CRHR2 in P15 family. Lower panel shows SNV of GRM8 in P4 family.

**Supplementary Figure 2.**
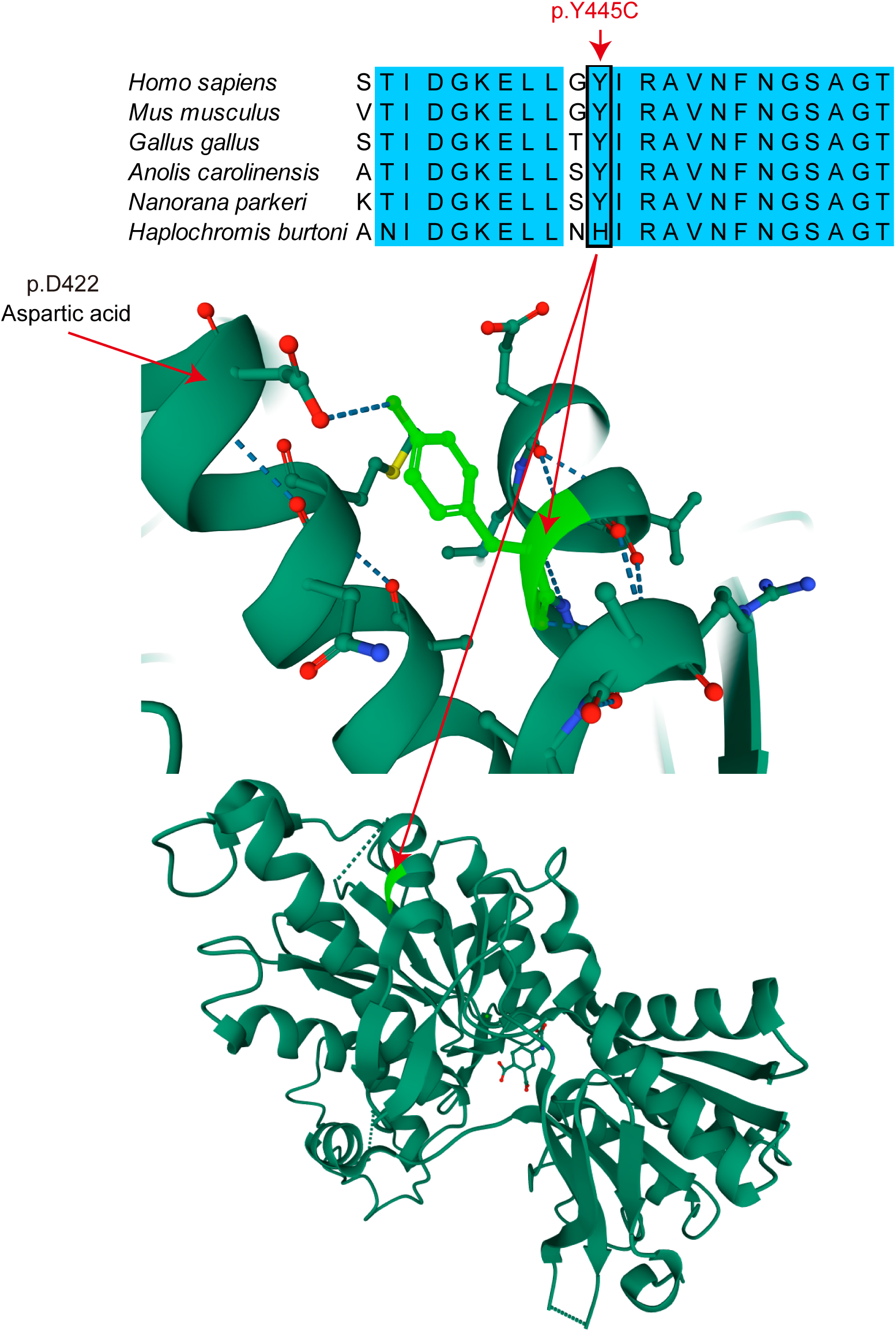
Multiple amino acid sequence alignment of GRM8 showing evolutionarily conserved amino acids and ATD structure of GRM8. Upper alignment indicates evolutionarily conserved amino acids. Black square indicates amino acid position corresponding to human Y445. Middle panel indicates ribbon diagram around Y445 (light green). Hydrogen bonds are depicted as blue dotted lines. Lower panel indicates ribbon representation for all ATDs including Y445 (light green). These ribbon diagram images were created using Mol* Viewer [1] with the PROTEIN DATA BANK (https://www.rcsb.org/) for PDB ID 6BSZ [2]. [1] D. Sehnal, S. Bittrich, M. Deshpande, et al., Mol* Viewer: modern web app for 3D visualization and analysis of large biomolecular structures, Nucleic acids research, 49 (2021) W431-w437. [2] Q. Chen, J.D. Ho, S. Ashok, et al., Structural Basis for (S)-3,4-Dicarboxyphenylglycine (DCPG) As a Potent and Subtype Selective Agonist of the mGlu(8) Receptor, Journal of medicinal chemistry, 61 (2018) 10040-10052.

**Supplementary Table1.**
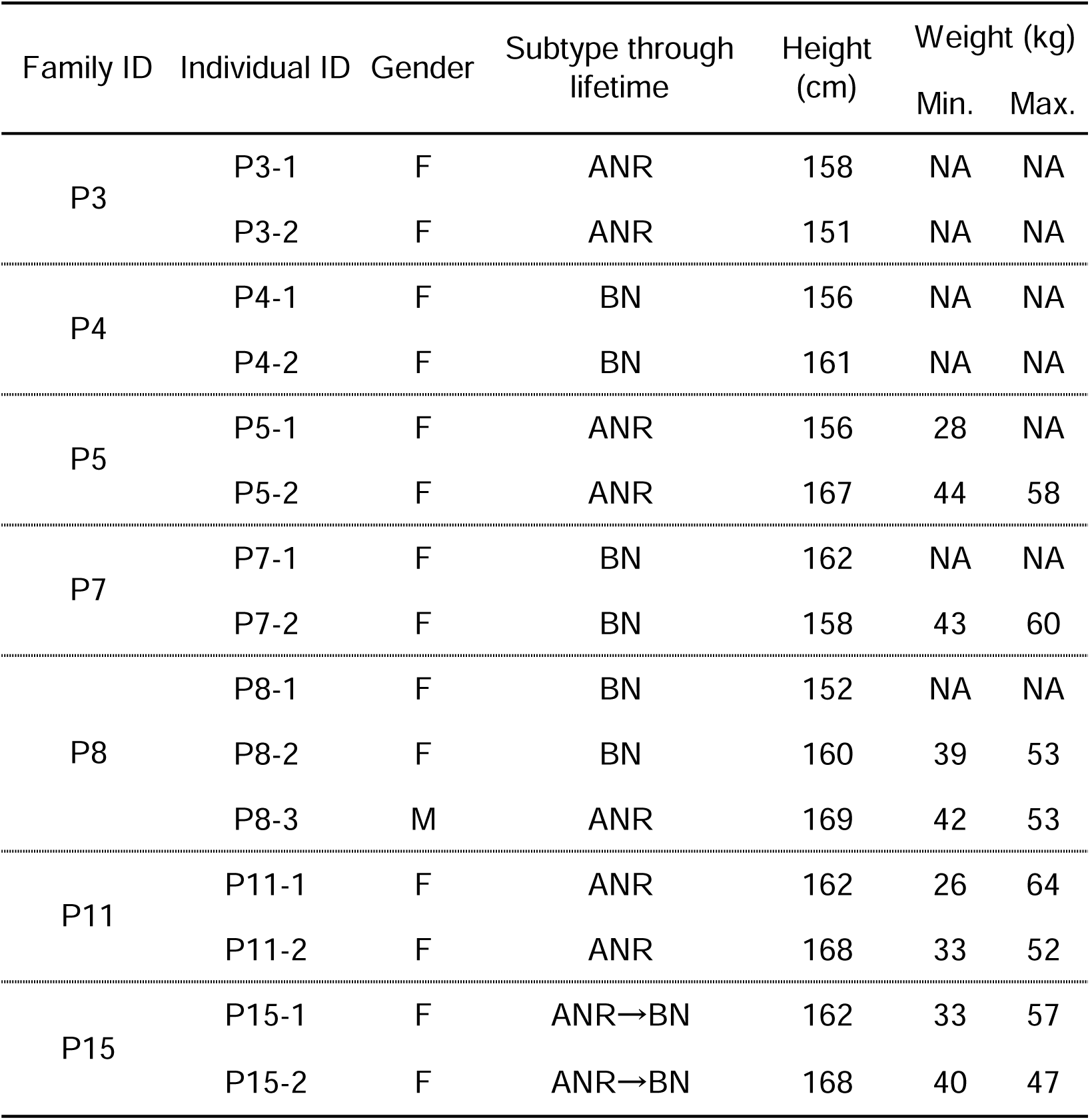
Clinical features of affected sib-pairs in this study.

| Family ID | Individual ID | Gender | Subtype through lifetime | Height (cm) | Weight (kg) |  |
| --- | --- | --- | --- | --- | --- | --- |
|  |  |  |  |  | Min. | Max. |
| P3 | P3-1 | F | ANR | 158 | NA | NA |
|  | P3-2 | F | ANR | 151 | NA | NA |
| P4 | P4-1 | F | BN | 156 | NA | NA |
|  | P4-2 | F | BN | 161 | NA | NA |
| P5 | P5-1 | F | ANR | 156 | 28 | NA |
|  | P5-2 | F | ANR | 167 | 44 | 58 |
| P7 | P7-1 | F | BN | 162 | NA | NA |
|  | P7-2 | F | BN | 158 | 43 | 60 |
| P8 | P8-1 | F | BN | 152 | NA | NA |
|  | P8-2 | F | BN | 160 | 39 | 53 |
|  | P8-3 | M | ANR | 169 | 42 | 53 |
| P11 | P11-1 | F | ANR | 162 | 26 | 64 |
|  | P11-2 | F | ANR | 168 | 33 | 52 |
| P15 | P15-1 | F | ANR→BN | 162 | 33 | 57 |
|  | P15-2 | F | ANR→BN | 168 | 40 | 47 |

**Supplementary Table2.**
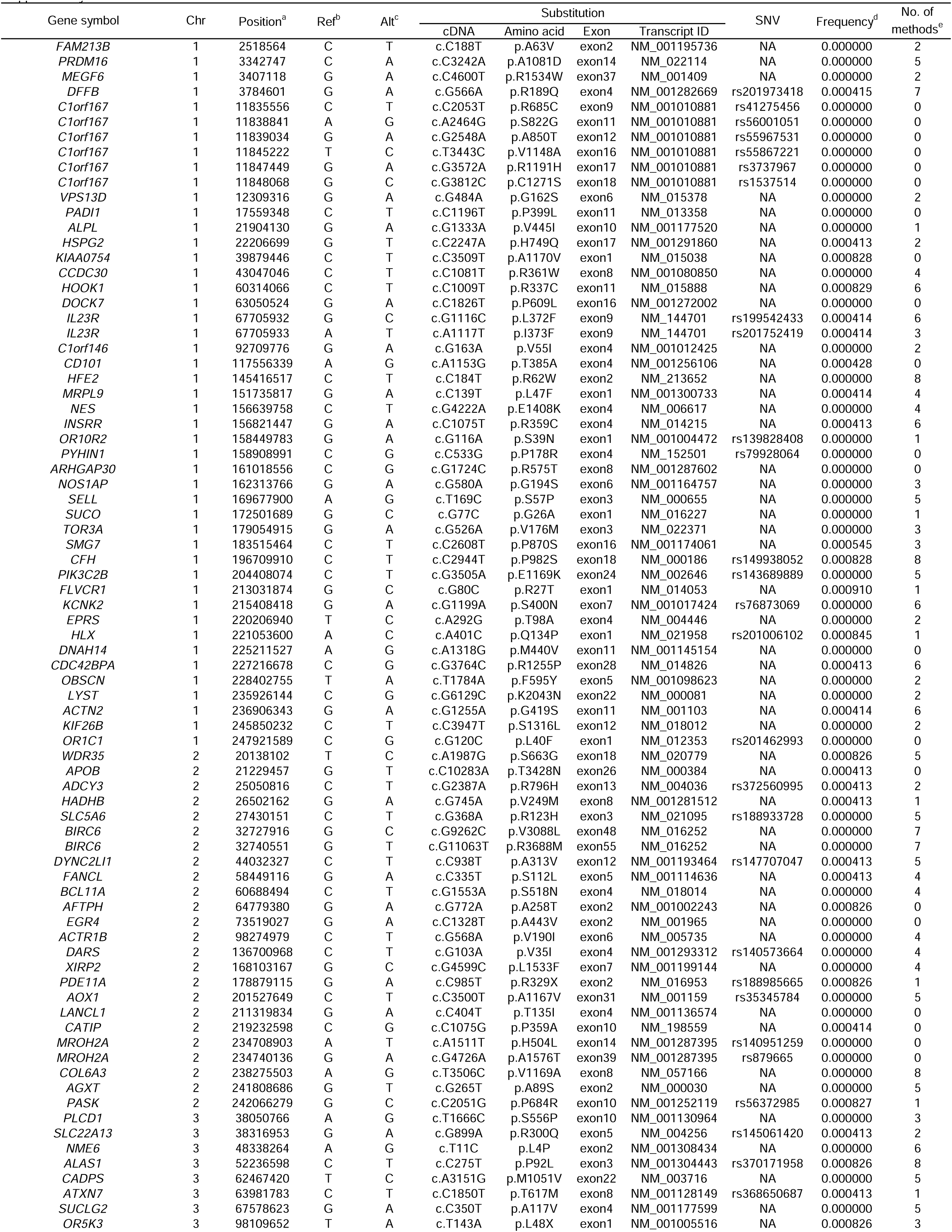
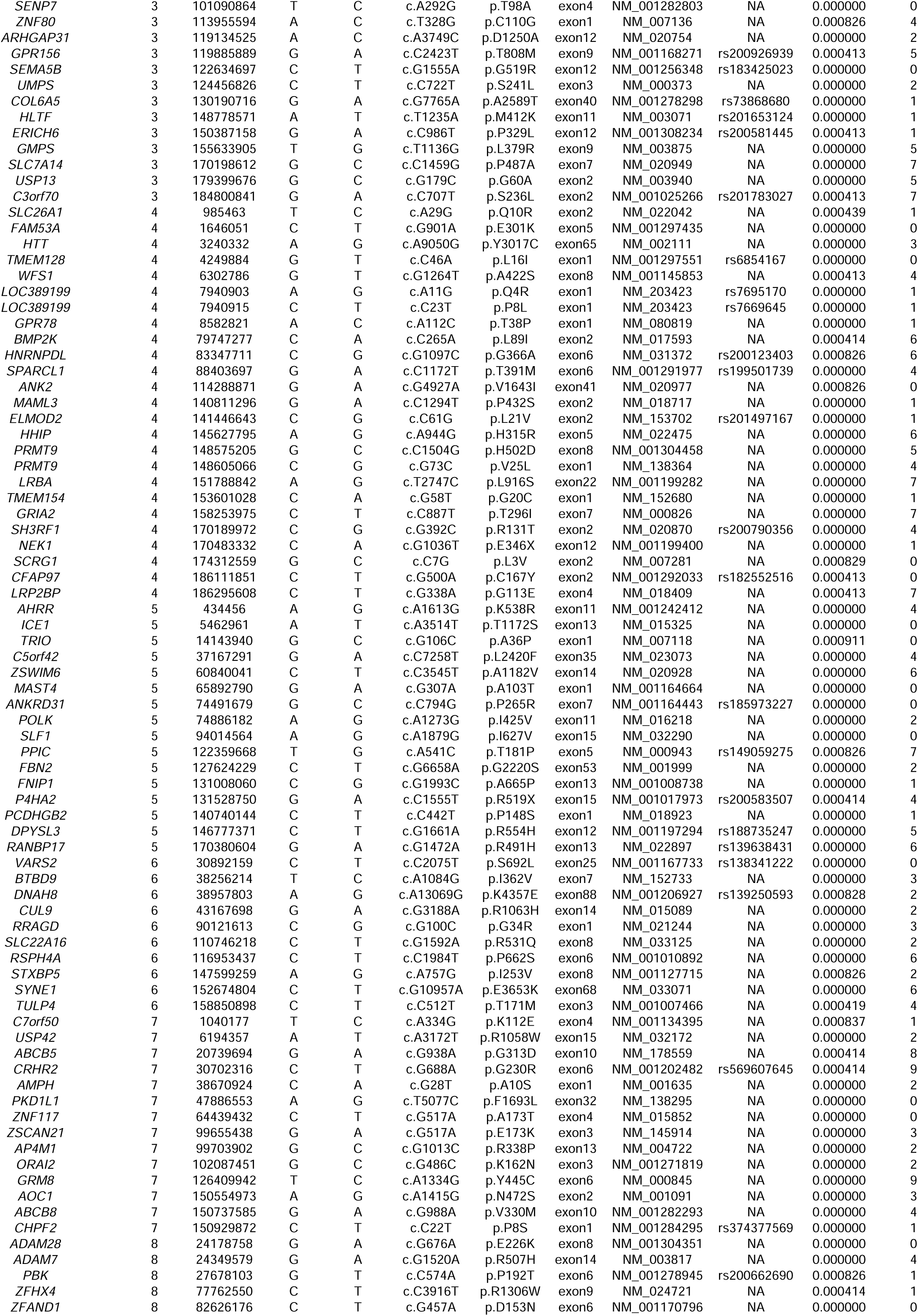
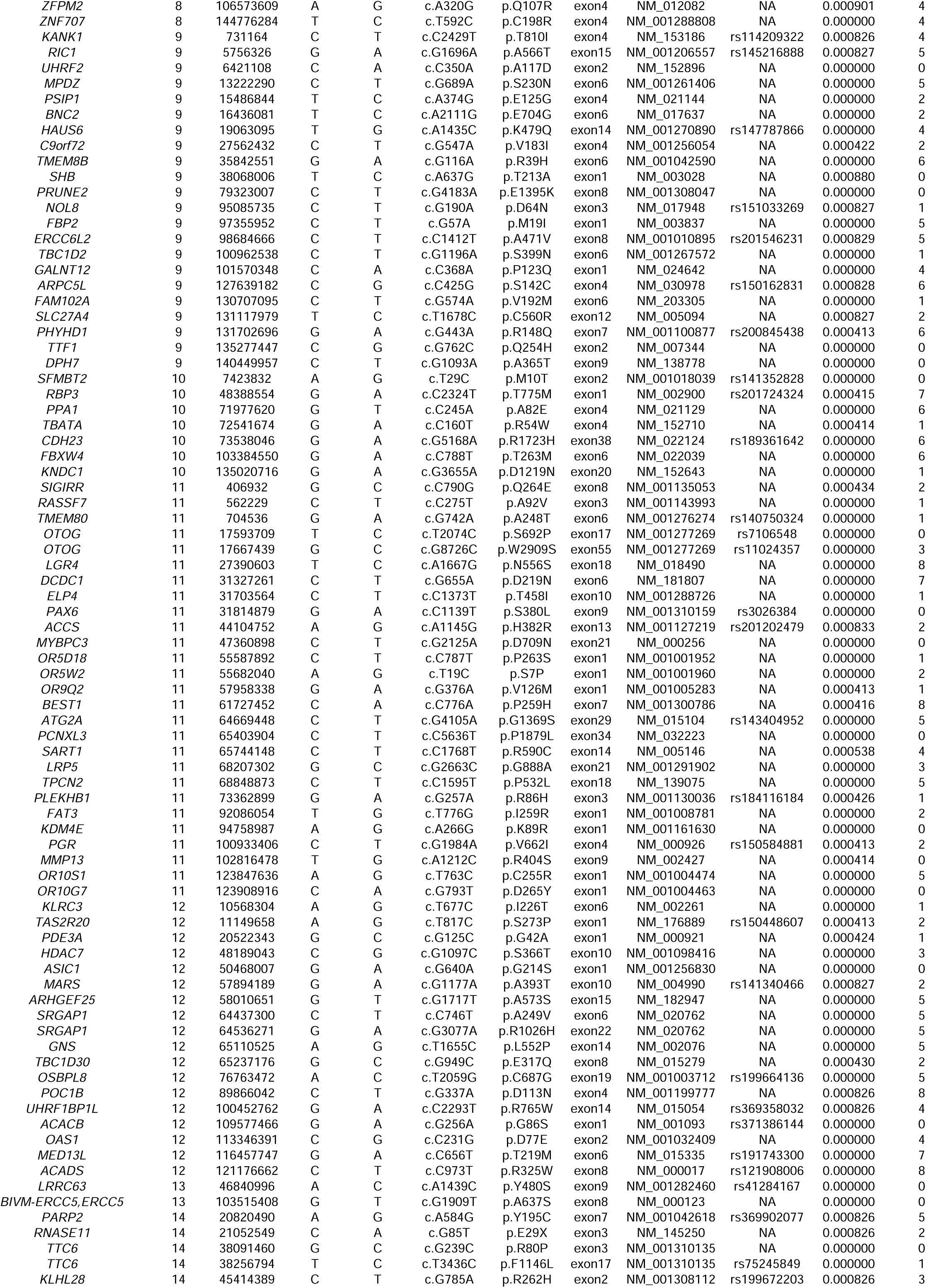
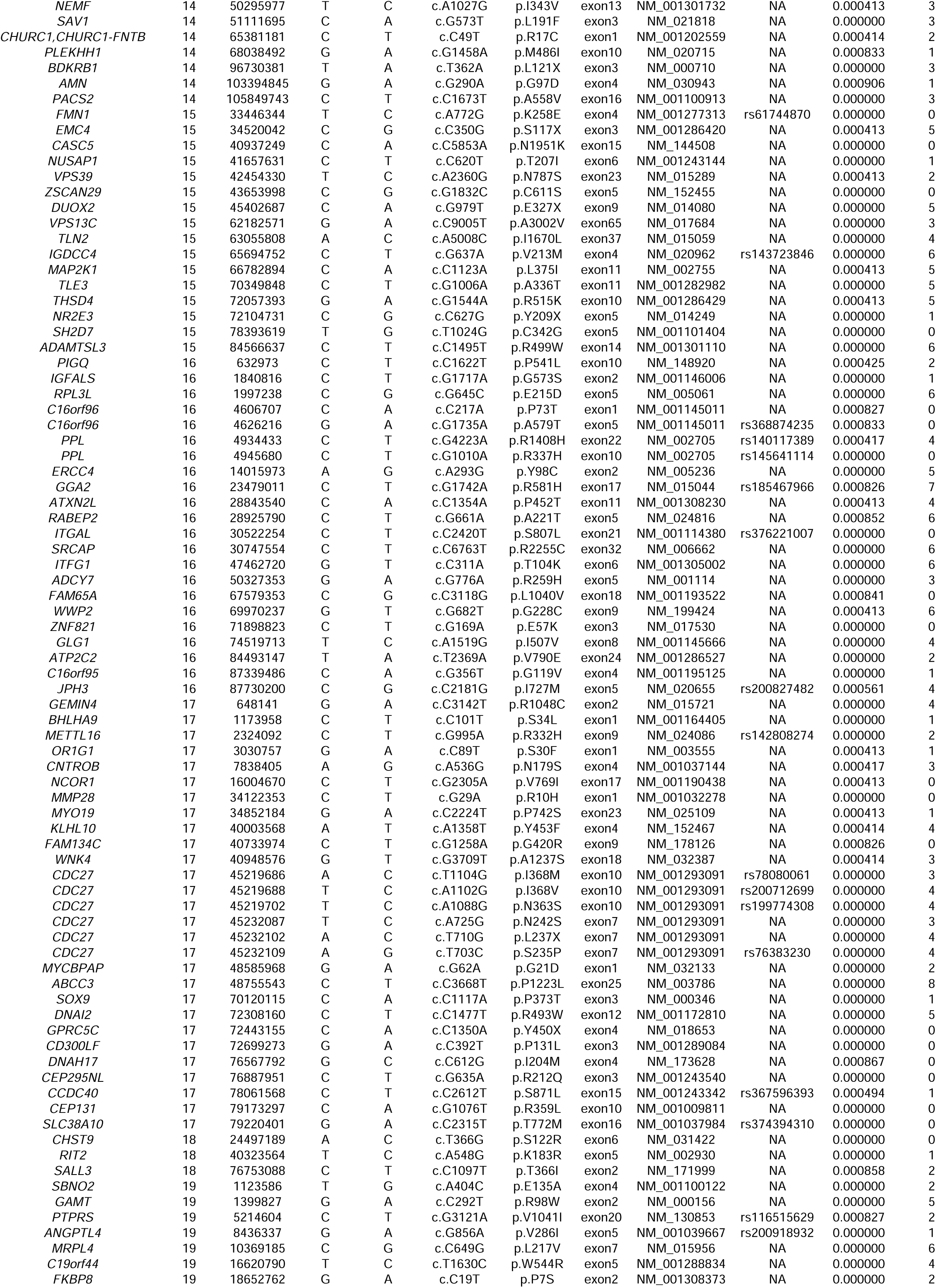
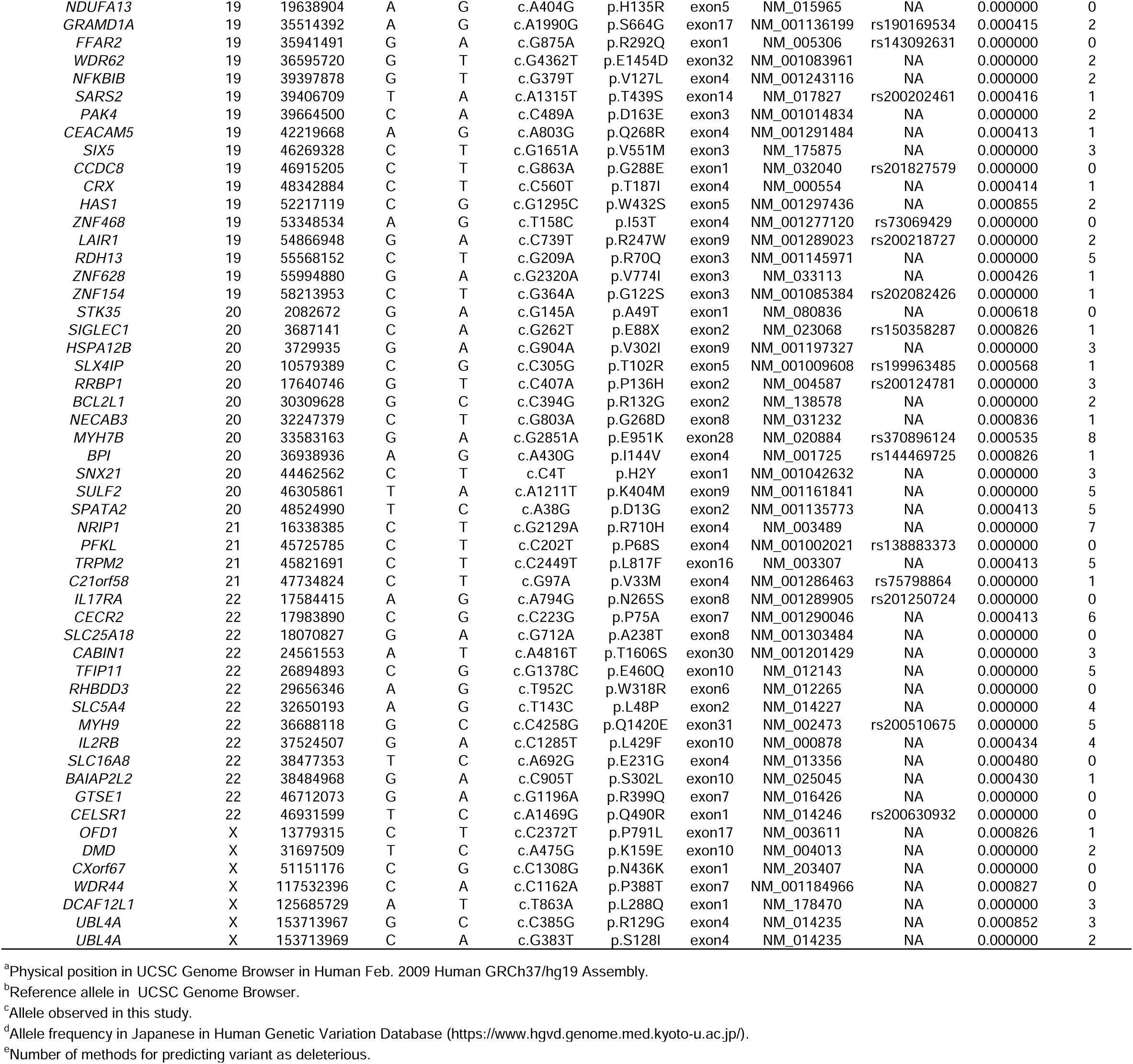

**Supplementary Table3.**
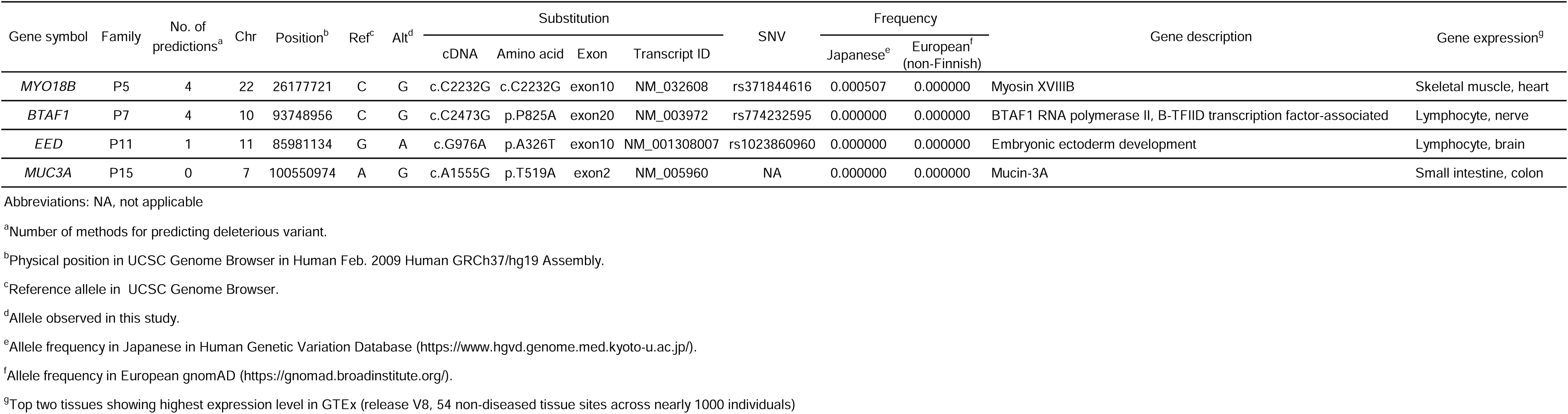
*De novo* variants in each family.

**Supplementary Table4.** Splice site variants in each family.

| Gene symbol | Family | Chr | Position<sup>a</sup> | Ref<sup>b</sup> | Alt<sup>c</sup> | Splice region | Substitution | SNV | Frequency |  | Gene description | Gene expression<sup>f</sup> |
| --- | --- | --- | --- | --- | --- | --- | --- | --- | --- | --- | --- | --- |
|  |  |  |  |  |  |  |  |  | Japanese<sup>d</sup> | European<sup>e</sup><br>(non-Finnish) |  |  |
| *ATG7* | P4 | 3 | 11468277 | G | C | splice acceptor | AG→AC | rs756690200 | 0.000000 | 0.000008 | Autophagy related 7 | Fibroblast, estist |
| *WDR45B* |  | 17 | 80579677 | T | G | splice acceptor | AG→CG | NA | 0.000000 | 0.000000 | WD repeat domain 45B | Testis, fallopian tube |
| *HBD* | P11 | 11 | 5255444 | C | T | splice acceptor | AG→AA | rs35408031 | 0.000000 | 0.000026 | Hemoglobin, delta | Whole blood, spleen |
| *LILRB3* |  | 19 | 54721187 | C | T | splice donor | GT→AT | rs147121453 | 0.000000 | 0.000134 | Leukocyte immunoglobulin-like receptor, subfamily B, member 3 | Whole blood, spleen |
| *FAM216B* | P15 | 13 | 43361020 | G | T | splice donor | GT→TT | NA | 0.000413 | 0.000000 | Family with sequence similarity 216, member B | Heart, fallopian tube |

Abbreviations: NA, not applicable
<sup>a</sup>Physical position in UCSC Genome Browser in Human Feb. 2009 Human GRCh37/hg19 Assembly.
<sup>b</sup>Reference allele in UCSC Genome Browser.
<sup>c</sup>Allele observed in this study.
<sup>d</sup>Allele frequency in Japanese in Human Genetic Variation Database (https://www.hgvd.genome.med.kyoto-u.ac.jp/).
<sup>e</sup>Allele frequency in European gnomAD (https://gnomad.broadinstitute.org/).
<sup>f</sup>Top two tissues showing highest expression level in GTEx (release V8, 54 non-diseased tissue sites across nearly 1000 individuals)

**Supplementary Table5.**
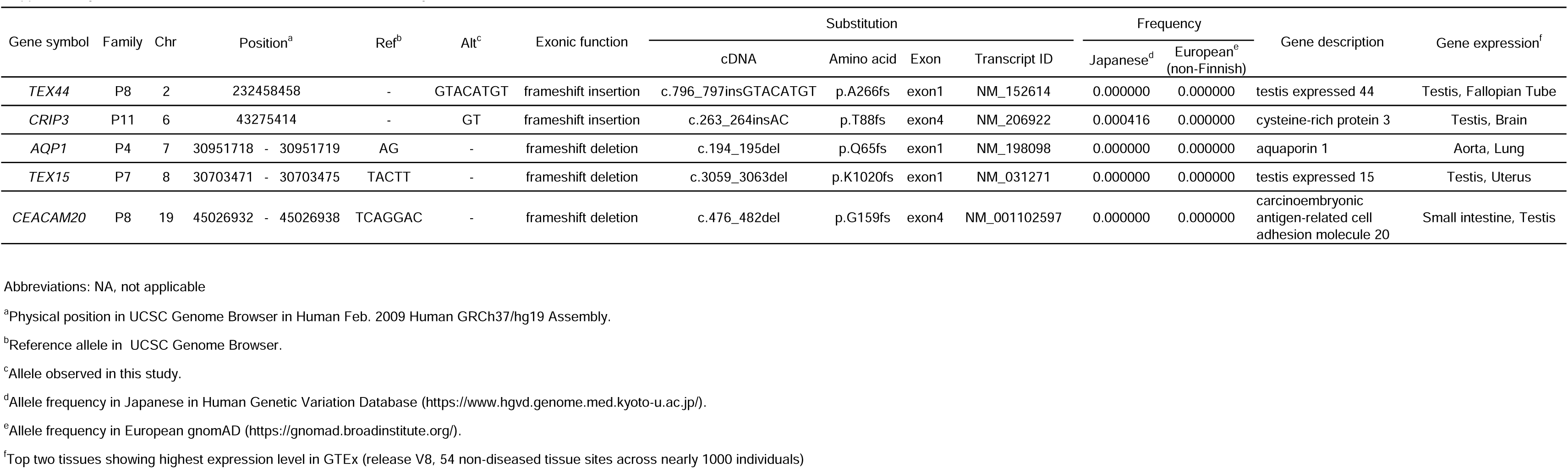
Insertion and deletion variants in each family.

